# Quantification of the tradeoff between test sensitivity and test frequency in COVID-19 epidemic - a multi-scale modeling approach

**DOI:** 10.1101/2021.02.15.21251791

**Authors:** Jonathan E. Forde, Stanca M. Ciupe

## Abstract

Control strategies that employ real time polymerase chain reaction (RT-PCR) tests for the diagnosis and surveillance of COVID-19 epidemic are inefficient in fighting the epidemic due to high cost, delays in obtaining results, and the need of specialized personnel and equipment for laboratory processing. Cheaper and faster alternatives, such as antigen and paper-strip tests, have been proposed. They return results rapidly, but have lower sensitivity thresholds for detecting virus. To quantify the effects of the tradeoffs between sensitivity, cost, testing frequency, and delay in test return on the overall course of an outbreak, we built a multi-scale immuno-epidemiological model that connects the virus profile of infected individuals with transmission and testing at the population level. We investigated various randomized testing strategies and found that, for fixed testing capacity, lower sensitivity tests with shorter return delays slightly flatten the daily incidence curve and delay the time to the peak daily incidence. However, compared with RT-PCR testing, they do not always reduce the cumulative case count at half a year into the outbreak. When testing frequency is increased to account for the lower cost of less sensitive tests, we observe a large reduction in cumulative case counts, from 57% to as low as 1.5% half a year into the outbreak and to 3.2% three years into the outbreak. The improvement is preserved even when the testing budget is reduced by one half or one third. Our results predict that surveillance testing that employs low-sensitivity tests at high frequency is an effective tool for epidemic control.

## 1 Introduction

Following the emergence of the novel coronavirus-2 severe acute respiratory syndrome (SARS-CoV-2) late in 2019 in Wuhan, China, the World Health Organization declared the COVID-19 pandemic on March 11, 2020. As of February 12, 2021, this pandemic has resulted in over 107.4 million confirmed infections and 2.3 million deaths worldwide [3].

Epidemiological data from nations such as South Korea, Iceland and Taiwan demonstrates that widespread surveillance using real time polymerase chain reaction (RT-PCR) testing, combined with contact tracing and quarantine measures, can be effective at limiting the spread of SARS-CoV-2 [23]. However, in many other nations, notably the United States, the testing infrastructure was insufficient to prevent viral spread.

Testing strategies across the world vary based on location, resources, and political considerations. While some countries test only symptomatic individuals, or those in need of hospitalization, others employed randomized testing early for surveillance and isolation [15]. In the United States, calls for frequent and widespread testing [34] have been associated with the reopening of the economy, schools and college campuses, and with the protection of essential workers [31, 32, 41].

Diagnosis of SARS-CoV-2 infection is usually achieved by RT-PCR nasopharyngeal test, considered the gold standard for SARS-CoV-2 detection. It has high (close to 100%) sensitivity in detecting active disease, but is expensive and can require up to 5 days of laboratory processing [37]. Moreover, it does not distinguish between transmissible and non transmissible infections. It has been proposed (but not yet been universally authorised by the FDA) that reporting virus titers from RT-PCR tests can help determine the stage of an individual’s disease [25]. For example, high viral RNA titer in the sample (above levels above 10^6^ virus RNA per ml) is considered a good proxy for infectiousness [17–19]. By contrast, low viral RNA that is still detectable in respiratory tracts and other specimens after disease resolution is believed to no longer be viable [24]. Quantifying virus RNA requires the recording the cycle threshold (Ct) value in the PR-PCR tests [13], with Ct< 30 being considered a threshold for infectivity [21]. RT-PCR’s viral limit of detection is around 10^2^ virus RNA per swab [10, 14, 38]. Detecting low virus RNA titers by the RT-PCR (corresponding to high Ct numbers, > 35) may not be relevant from an epidemiological point of view, since they are associated with fewer tissue-culture infective viral particles [13, 35] and, hence, low probability of transmission [12]. Therefore less sensitive tests that are easier to use and give instantaneous results may be a good substitute for RT-PCR [25].

Several fast detection antigen and molecular strategies have obtained emergency use authorization (EUA) from FDA. They have the potential of more quickly detecting and isolating symptomatic and asymptomatic infections compared to laboratory-based diagnostic methods [2]. Among them are rapid antigen tests, such as the Abbott pharmaceuticals’ BinaxNOW^™^ [1, 33], serological tests [16], and hypothesized but not yet manufactured at-home paper-strip tests [22,25]. Such tests, however, have lower overall sensitivities and only detect higher virus titers [21]. For example, the BinaxNOW^™^ antigen rapid test has sensitivity levels of 85.7% for Ct< 25 (when the infection is still transmissible), and 36.4% for Ct> 30 (when the infection may no longer be transmissible) [22], making it an acceptable alternative to RT-PCR, especially since they are cheaper and produce results quickly, in as little as 15 minutes [1, 16]. Frequent testing with cheap, low-sensitivity tests, may therefore be beneficial for population surveillance and quarantining practices [22, 25].

An example of the tradeoff between a single RT-PCR test and multiple low-sensitivity alternate tests is given in Figure 1. In one scenario, an RT-PCR test is administrated for diagnosis of a symptomatic individual, who has been infectious (defined as virus above 10^6^ virus RNA per ml [17–19]) and transmitting the virus several days before test administration and 1-2 days before symptoms onset [18] (see Figure 1, panel A, red circle). In a second scenario, a surveillance test is administered to an asymptomatic individual, who may or may not be transmitting the virus at the time of testing (see Figure 1, panel B, red circle). In a third scenario, a cheaper, less sensitive test is administrated repeatedly in the same patient (see Figure 1, panel C, yellow circles). The test fails to identify a positive case at the time of infectiousness due to sensitivity issues, but still has the potential of discovering and reporting the infection earlier in the individual’s transmissibility window, compared to the RT-PCR (see Figure 1, panel C, fourth yellow circle). Determining when such a tradeoff occurs, and how frequent the low-sensitivity tests should be administrated in order to outperform RT-PCR, is important for designing interventions.

**Figure 1:**
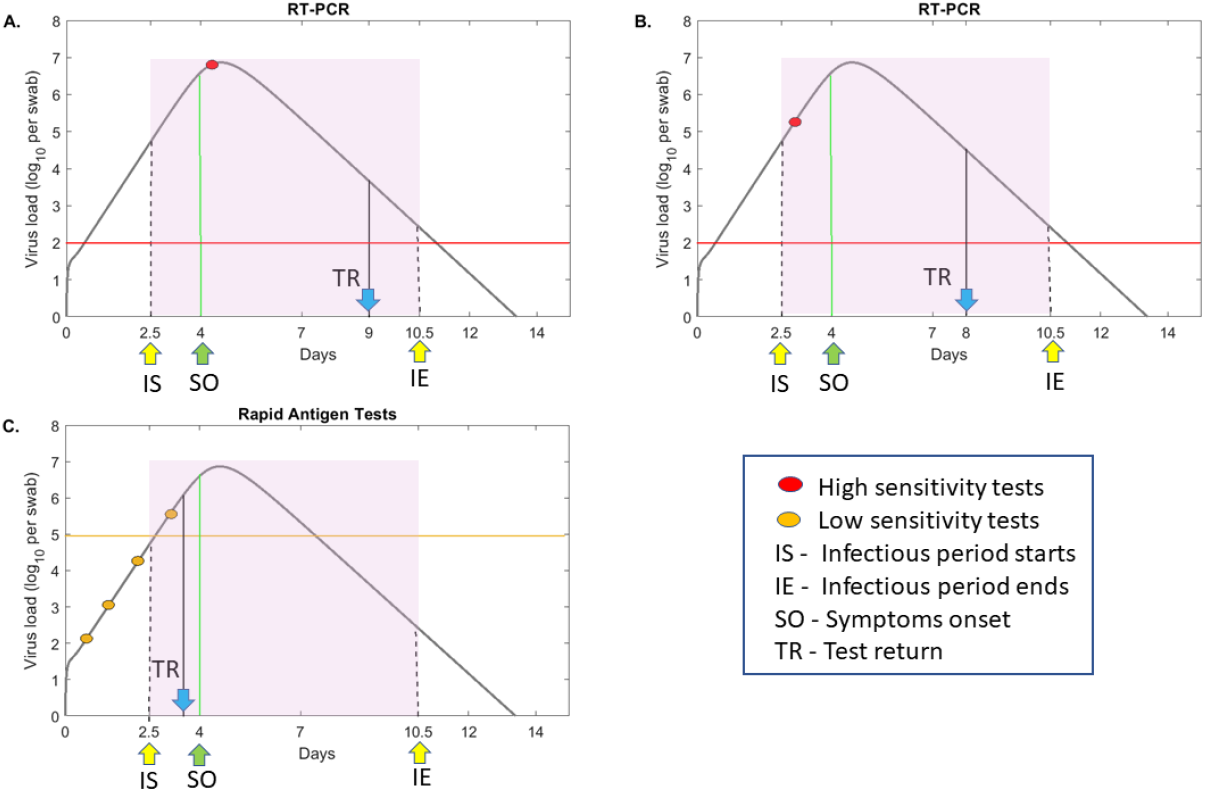
RT-PCR versus rapid testing practices. log_10_ virus load per swab over time as given by model (1) (grey curves) for values in [20]. Patients are assumed to be infectious from *t* = 2.5 days (IS) till *t* = 10.5 days (IE) (shaded region) and symptomatic beginning on day *t* = 4 (SO). Panels A and B depict testing with a high-sensitivity RT-PCR test with detection threshold log_10_(*V*) = 2 per swab (red line) and test return delay of five days. In panel A, the test occurs immediately following symptoms onset, and in panel B, the test occurs before symptoms onset (red circles). Panel C depicts frequent testing (yellow circles) with a low-sensitivity test with detection threshold log_10_(*V*) = 5 per swab (yellow line) and test return delay of one half day. TR shows the time of positive test result.

In this study, we develop a multi-scale within-host between-host time-since-outbreak model and investigate its dynamics under testing. The within-host model gives information on the time of infectiousness onset and the time interval when a test detects an infection by looking at the virus dynamics inside an infected individual. The between-host model connects these events with transmission at the population level. We investigate testing strategies with assays of different sensitivities, frequencies, and delays in test returns. We will deem optimal a testing strategy that flattens the infection curve best, under either the same testing frequency or the same monetary cost.

## 2 Methods

### 2.1 Within-host model

To generate within-host virus profiles, we use the target cell limitation model of SARS-CoV-2 kinetics developed by Ke *et al*. [20], which was fitted to virus levels measured in pharyngeal swabs and sputum samples of patients infected through contact with the same index case [10, 39]. Briefly, the model considers the interaction between uninfected epithelial cells, *T*_*j*_; exposed epithelial cells, *E*_*j*_; infected epithelial cells, *I*_*j*_; and virus, *V*_*j*_ in upper (URT) and lower (LRT) respiratory tracts *j* ∈ {1, 2}, as in other acute infections [4–7, 9, 28, 29, 36]. Target cells in each tract are infected at rates *β*_*j*_, exposed cells become infectious at rates *k*_*j*_, and infected cells produce new virions at rates *p*_*j*_. Infected cells die at rates *δ*_*j*_ and virus particles are cleared at rate *c*, independent of the tract. The two tracts are linked via the virus populations, with a proportion *g*_12_ of *V*_1_ migrating from URT to LRT and a proportion *g*_21_ of *V*_2_ migrating from LRT to URT. The model describing these interactions is given by

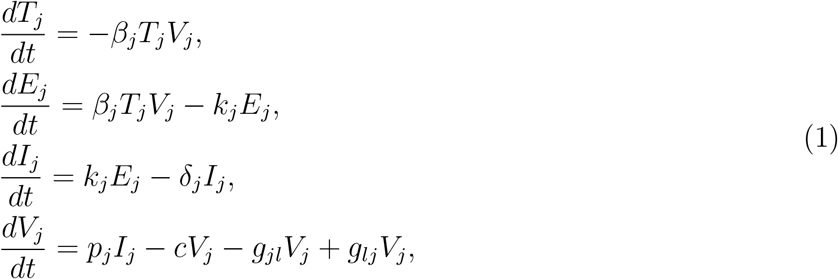

where *j* ≠ *l* ∈ {1, 2}. Ke *et al*. [20] assumed that the pharyngeal swabs data *V*_*T*_ and sputum data *V*_*S*_ in [10, 39] are proportional to the predicted URT and LRT virus loads given by model (1), *V*_*T*_ = *f*_1_*V*_1_ and *V*_*S*_ = *f*_2_*V*_2_. They assumed that parameters {*k*_*j*_, *c, g*_21_} are known and fitted the remaining parameters {*β*_*jT*_, *δ*_*j*_, *π*_*j*_, Γ} to the data, where *β*_*jT*_ = *β*_*j*_*/f*_*j*_, *π*_*j*_ = *f*_*j*_*p*_*j*_ and Γ = *f*_2_*g*_12_*/f*_2_. Here, we use the estimates from one case in Ke *et al*. (patient E) to generate the virus profiles *V* = *V*_*T*_ = *f*_1_*V*_1_ which we use in the multi-scale transmission model eqs. (5) and (3). A summary of the parameters used in eq. (1) is given in Table 1.

**Table 1:** Parameter values and initial conditions used in model (1)

| Fixed parameters | Description | Value | Source |
| --- | --- | --- | --- |
| $k_j$ | Eclipse phase duration | 4/day | [30] |
| $c$ | Viral clearance | 10/day | [4] |
| $g_{21}$ | Transport between tracts | 0 | [20] |
| Estimated parameters | Description | Value | Source |
| $\beta_{1T}$ | Infection rate in URT | $5.1 \times 10^{-7}/\text{swab} \times \text{day}$ | [20] |
| $\beta_{2T}$ | Infection rate in LRT | $7 \times 10^{-7}/\text{ml} \times \text{day}$ | [20] |
| $\pi_1$ | URT virus production | 50/day | [20] |
| $\pi_2$ | LRT virus production | 0.34/day | [20] |
| $\delta_1$ | URT cell death | 2/day | [20] |
| $\delta_2$ | LRT cell death | 0.53/day | [20] |
| $\Gamma$ | - | 0.01 | [20] |
| Initial conditions | Description | Value | Source |
| $T_1(0)$ | Epithelial cells in URT | $4 \times 10^6/\text{ml}$ | [20] |
| $T_2(0)$ | Epithelial cells in LRT | $4 \times 10^8/\text{ml}$ | [4] |
| $E_j(0)$ | Exposed epithelial cells | 0 | [20] |
| $I_1(0)$ | Infectious epithelial cells in URT | 10 | [20] |
| $I_2(0)$ | Infectious epithelial cells in LRT | 1 | [20] |
| $V_j(0)$ | Virus | 0 | [20] |

### 2.2 Between-host model

We model the interaction between a susceptible class *S*(*t*), infected class of asymptomatic individuals, *i*_*a*_(*τ, t*), and infected class of symptomatic individuals, *i*_*s*_(*τ, t*). The independent variables are *τ*, the age of infection in an individual, and *t*, the time-since-outbreak in the population. We assume the individual infection status is given by its virus profile at time *τ, V* (*τ*), with *V* (*τ*) = *V*_*T*_ (*τ*) = *f*_1_*V*_1_(*τ*) being the solution of system eq. (1). We assume that *β* is the transmission rate, *λ*_*j*_ the force of infection, *b* the birth rate, *µ* the death rate, *m*_*j*_ the disease induced mortality rates, and *j* ∈ *{a, s}*. In the absence of testing, the model is given by

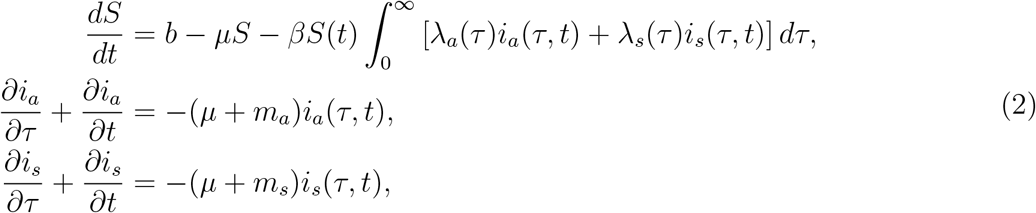

for 0 ≤ *τ* ≤ 14. For *τ* > 14, infections are considered to be resolved, and recovered individuals are not susceptible to reinfection. The boundary and initial conditions are

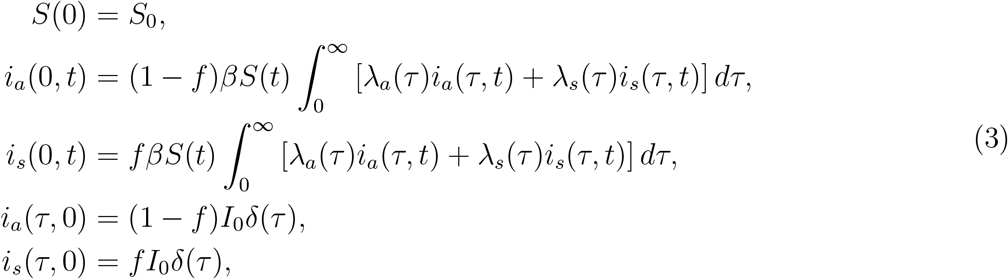

where *f* is the fraction of infections that are symptomatic. Parameters {*β, µ, m_a_, m_s_, f*} are taken from literature, and *δ*(*τ*) is the Dirac delta function. A summary of the parameters we used in eqs. (2) and (3) is given in Table 2.

**Table 2:** Parameter values and initial conditions used in model eq. (5).

| Fixed parameters | Description | Value | Source |
| --- | --- | --- | --- |
| $\beta$ | Transmission rate | 0.25/day | |
| $b$ | Birth rate | $7 \times 10^{-4}/\text{day}$ | [27] |
| $\mu$ | Death rate | $7 \times 10^{-4}/\text{day}$ | [27] |
| $m_j$ | Disease induced mortality rate | $10^{-3}/\text{day}$ | |
| $f$ | Fraction of symptomatic infections | 0.7 | [17] |
| $\gamma$ | Relative asymp. infectiousness | 0.7 | |
| $\ell$ | Test return delay | varied | |
| $\tau_j^1$ | Age of onset of virus detectability | varied (days) | |
| $\tau_j^2$ | Age of onset of infectiousness | 2.5 days | [39] |
| $\tau_j^3$ | Age of end of infectiousness | 10.5 days | [17, 18] |
| $\tau_j^4$ | Age of loss of virus detectability | varied (days) | |
| Initial conditions | Description | Value | Source |
| $S(0)$ | Susceptible population | 0.99 | |
| $i_s(\tau, 0)$ | Infected symptomatic population | $0.01f\delta(\tau)$ | |
| $i_a(\tau, 0)$ | Infected asymptomatic population | $0.01(1 - f)\delta(\tau)$ | |

### 2.3 Daily testing rate

We determine a per capita random testing rate, *ρ*_rand_, corresponding to an overall testing capacity of *C* tests per day, as follows. If subjects are removed from a population *P* by testing at per capital rate *ρ*_rand_, then the remaining untested population is given by

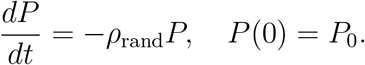

The total number of tests administered in a given day is 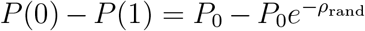. Setting this equal to the testing capacity *C*, we find that the daily random testing rate corresponding to the administration of *C* test is given by

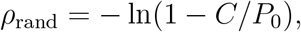

so long as *C* < *P*_0_. Thus, if *N* (*t*) is the population subject to random testing at time *t*, the time-dependent continuous testing rate is

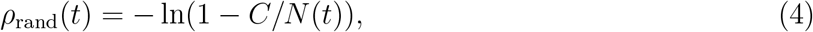

for *N* (*t*) < *C*.

### 2.4 Between-host model with testing

Given virus profiles for infected individuals, we link test sensitivity to the ages of infection during which virus load is above the sensitivity threshold. Similarly, we determine the ages of infection during which the virus load is high enough to allow transmission. We define

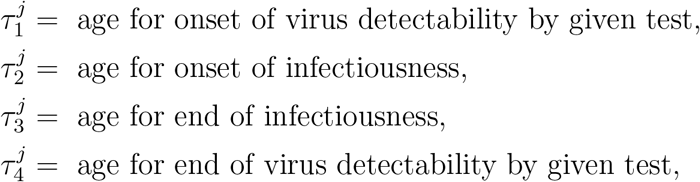

where *j* ∈ {*a, s*}. The force of infection functions *λ*_*j*_ are

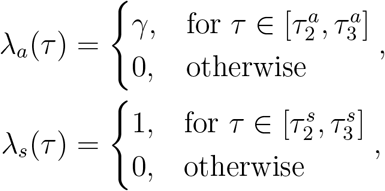

where parameter 0 < *γ* < 1 represents the relative infectiousness of asymptomatic carriers, in comparison with symptomatic carriers. The case detection rate functions *r*_*j*_(*τ, t*) become

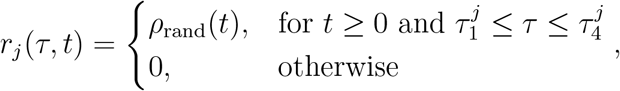

where *j* ∈ {*a, s*}. We assume a test return delay of *ℓ* days, and that individuals who receive a positive test result are isolated, and can no longer transmit the virus. Lastly, we ignore the possibility of reinfection. The between-host model equations under testing become

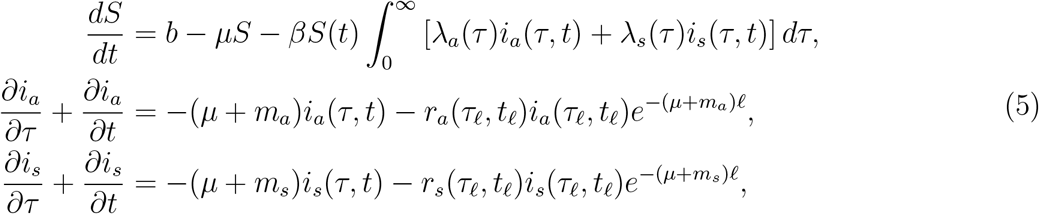

where *τ*_*ℓ*_ = *τ* − *ℓ* and *t*_*ℓ*_ = *t* − *ℓ*. The cumulative number of cases at time *t*, Σ(*t*), is given by the equation

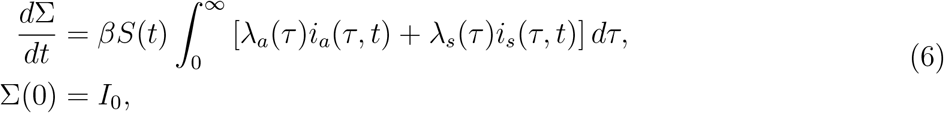

and the cumulative number of detections at time *t, P* (*t*), is given by the equation

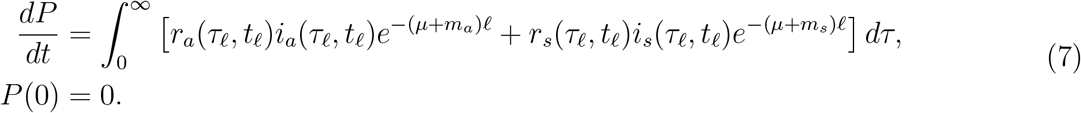

The boundary and initial conditions (see eq. (3)) and parameters {*b, β, µ, m_a_, m_s_, f*} (see Table 2) are as before. The return delay *ℓ* and ages 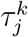 for *j* ∈ {*a, s}* and *k* ∈ {1,.., 4} vary among tests. A summary of parameters and initial conditions are given in Table 2 and the integration method is described in the Appendix.

## 3 Results

### 3.1 The relationship between test sensitivity and virus titers

We connect test sensitivities, defined as the threshold above which a test is able to correctly identify a true positive COVID-19 case, to the times in an individual’s infection when the SARS-CoV-2 titers are above this threshold. Temporal virus titers are determined using a previously published mathematical model of within-host virus dynamics [20]. The model, given by eq. (1), assumes interactions between target, exposed and infectious epithelial cells and SARS-CoV-2 in the upper and lower respiratory tracts, which are connected by virus shedding (Methods). A fraction of the upper respiratory tract virus, *V*_*T*_ = *f*_1_*V*_1_ in model eq. (1), was fitted [20] to longitudinal SARS-CoV-2 data in swab samples from mild infections [10, 39]. Here, we use the parameter estimates of one infected individual (patient E in [20]) to determine a generic theoretical curve for the SARS-CoV-2 levels over time (see Figure 1, grey curves). The viral curve spans over several milestones in an individual’s infection: infectiousness period (Figure 1, shaded region); symptoms onset (Figure 1, green arrow) and the infectiousness status at the time of test return (Figure 1, blue arrow). Under this viral motif, we find that a RT-PCR test that detects 10^2^ virus RNA per swab will be able to detect virus in a sample taken as early as 13 hours and as late as 10.9 days post-infection. This interval is longer than the infectiousness period of eight days (2.5 to 10.5 days post-infection). By contrast, a rapid test, such as the Abbott Pharmaceuticals’ BinaxNOW^™^ antigen rapid test [1] which can detect 10^5^ virus RNA per swab, will be able to detect virus in a sample taken as early as 2.7 days and as late as 7.4 days post-infection, an interval 3.3 days shorter than the infectiousness period. Lastly, a low-sensitivity test that detects 10^6^ virus RNA per swab, will be able to detect virus in a sample taken as early as 3.5 days and as late as 6.2 days post-infection, an interval 5.1 days shorter than the infectiousness period. We are interested in determining when the decrease in sensitivity can be compensated by increased testing frequency and/or reduced time in test return.

### 3.2 Mathematical model of testing during SARS-CoV-2 transmission

We develop a between-host SI model for a well-mixed population, given by a system of ordinary and partial differential equations. It considers the interactions between susceptible individuals, *S*(*t*), and two types of infected individuals: asymptomatic, *i*_*a*_(*τ, t*), and symptomatic, *i*_*s*_(*τ, t*). The independent variables are the age of infection in an individual, *τ*, and the time-since-outbreak in the population, *t* (see model eqs. (2) and (3) in Methods). We set an individual’s incubation period to the previously estimated value of 4 days (patient E in [39]); and assume that infectiousness occurs 1.5 days before the symptoms onset, *τ*_2_ = 2.5 days [18], and ends eight days later, *τ*_3_ = 10.5 days.

### 3.3 Quantifying the tradeoff between test sensitivity and return delay

To determine the effect on the total population, 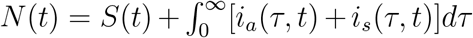, of tests with different sensitivities, frequencies, and return delays, we expand the SI model to include the age of infection at which a test first gives a positive result, *τ*_1_; the age of infection past which a test can no longer detect the virus, *τ*_4_; the return delay, *ℓ*; and the daily testing capacity, *C*. Assuming that surveillance testing occurs in a randomized manner, we calculate a continuous testing rate *ρ*_rand_(*t*), which is equal to −ln(1 − *C/N* (*t*)). This connects the daily testing capacity *C* with the population that is subject to random testing on a particular day, *N* (*t*) (see eq. (4) in Methods, for a derivation). The resulting system of differential equations (see model eqs. (5) and (3) in Methods) was used to predict epidemic outcomes under three testing regimes: an RT-PCR test, a rapid antigen test, and a paper-strip test. We assume a fixed daily testing capacity of 10% of the initial population, *C* = 0.1, which is administrated randomly among the groups, and an initial 1% of the population being infected. A portion *f* = 0.7 of the initial infected population is in the symptomatic class, and the remaining 1 − *f* = 0.3 is in the asymptomatic class.

Under the RT-PCR test with fixed *C* = 0.1 daily testing capacity rate, detection interval (*τ*_1_, *τ*_4_) = (0.55, 10.95) days, and delay in test results of *ℓ* = 5 days, model eq. (5) predicts a peak infection 42 days after the start of the outbreak, when 8.4% and 3.6% of the population have symptomatic and asymptomatic infections, respectively (see Figure 2, panel A, red and blue curves). When we ignore reinfection of recovered individuals, the infection dies out 75 days after the start of the outbreak, when less than 0.1% of the population is infected. A total of 57% of the population had the disease half a year into the outbreak, and the test successfully detected 96.4% of these (see Figure 2, panel A, magenta versus green curves). The highest daily incidence of 1.27% occurs 39 days after the start of the outbreak (see Figure 2, panel A, yellow bars). The daily detection rates lagged due to test return delays, peaking 49 days after the start of the outbreak (see Figure 2, panel A, blue bars).

**Figure 2:**
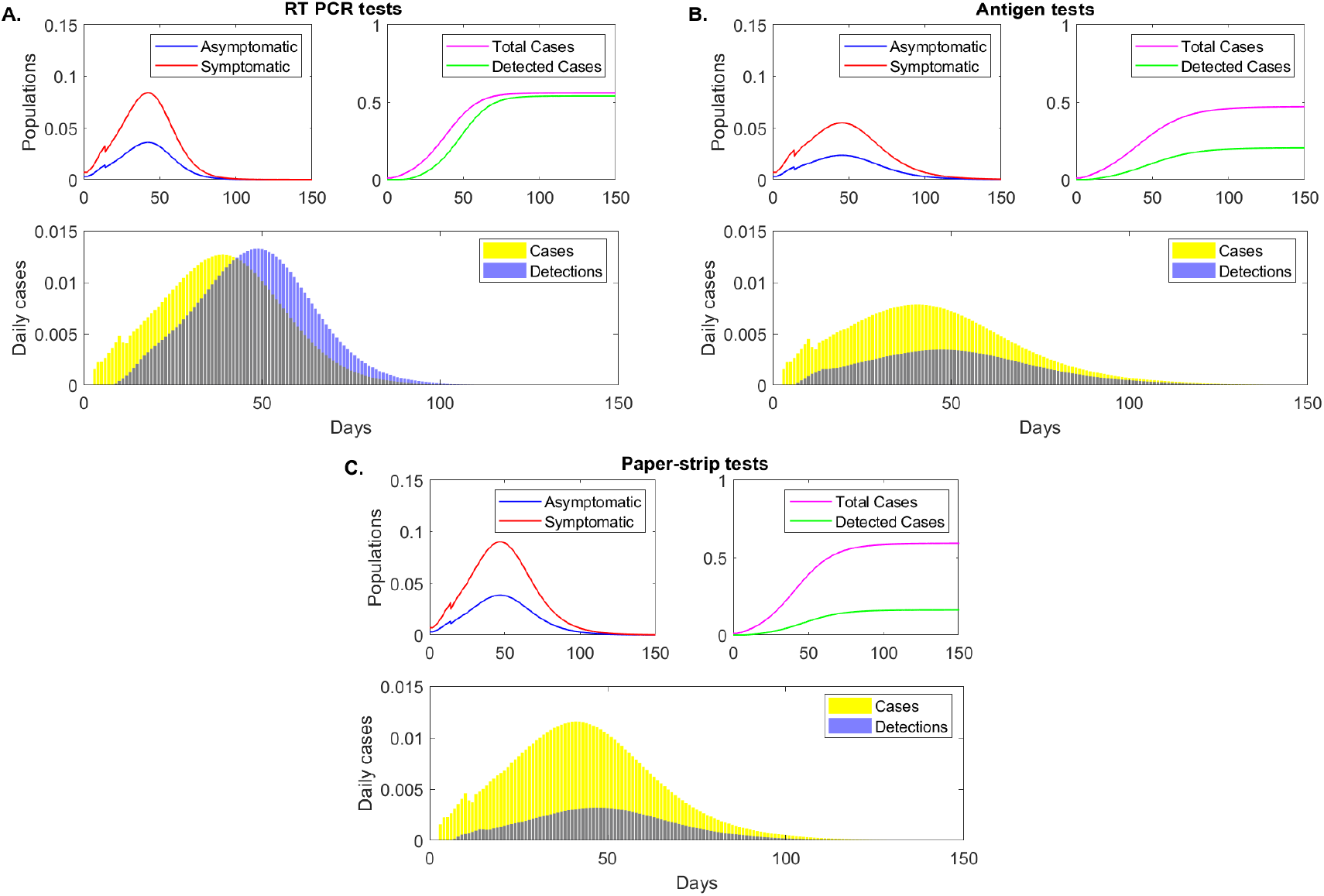
Epidemic dynamics over time. Sample epidemic dynamics results from varying testing regimes, as given by model eq. (5) for fixed testing capacity. Panel A: RT-PCR, detection threshold log_10_(*V*) = 2, test return delay 5 days; Panel B: antigen test, detection threshold log_10_(*V*) = 5, test return delay 0.5 days; Panel C: paper-strip test, detection threshold log_10_(*V*) = 6, test return delay 0.1 days. Upper left figures: asymptomatic (blue), symptomatic (red) populations over time. Upper right figures: cumulative positive cases (magenta) and cumulative detected cases (green) over time. Lower figures: daily new cases (yellow bars) and daily new case detections (blue bars).

Under a rapid antigen test with fixed *C* = 0.1 daily testing capacity rate, detection interval (*τ*_1_, *τ*_4_) = (2.77, 7.37) days, and delay in test results of *ℓ* = 0.5 days, model (5) predicts a peak infection at 45 days after the start of the outbreak, three days later than in the RT-PCR case. At that time, 5.5% and 2.4% of the population have symptomatic and asymptomatic infections, respectively, lower than in the RT-PCR testing scenario (see Figure 2, panel B, red and blue curves). While the infection does not decay to less than 0.1% daily case until day 90 after the start of the outbreak, the total population infected half a year after the start of the outbreak is 47%, lower than in the RT-PCR case where 57% individuals had the infection. This occurs in spite of only 42.6% of infections being detected (see Figure 2, panel B, magenta versus green curves). The highest daily incidence of 0.78% occurs 40 days after the start of the outbreak (see Figure 2, panel B, yellow bars) and daily detection rates peak 8 days later, at day 48 (see Figure 2, panel B, blue bars).

Lastly, under an even faster yet lower sensitivity paper-strip (or antigen) test with fixed *C* = 0.1 daily testing capacity rate, detection interval (*τ*_1_, *τ*_4_) = (3.48, 6.14) days, and delay in test results of *ℓ* = 0.1 days, model eq. (5) predicts a peak infection 47 days after the start of the outbreak, when 9% and 3.8% of the population have symptomatic and asymptomatic infections, respectively (see Figure 2, panel C, red and blue curves). While the peak of infection is delayed, the daily infections are higher than both those in the RT-PCR and antigen testing approaches. At half a year after the start of the outbreak 59% of population has been infected. Of those, 27.1% have been detected (see Figure 2, panel C, magenta versus green curves). The highest daily incidence of 1.15% occurs 41 days after the start of the outbreak (see Figure 2, panel C, yellow bars), lower than in the RT-PCR but higher than in the antigen testing approach. The daily detection rates peak 46 days after the start of the outbreak (see Figure 2, panel C, blue bars).

These results show that, with fixed testing capacity, tests that return results quickly, slightly flatten the daily incidence curve. The sensitivity is important, however, with low-sensitivity (corresponding to rapid antigen tests) resulting in a slight reduction in the total infections half a year into the outbreak, and super-low-sensitivity (corresponding to paper-strip tests) resulting in increased total infections. To more closely determine the relationship between the total cases half a year after the start of the outbreak, the return delays, and the test sensitivities, we derive a heat map for smaller sensitivity and delay increments (see Figure 3, panel A). We find that the RT-PCR holds better results than a test that detects 10^3^, 10^4^ and 10^5^ RNA per swab in half a day, only when the return is shorter than 2, 2.8 and 4.2 days, respectively. This means that, under the same daily test capacity, low-sensitivity tests can be a preferable surveillance resource in areas where there are long delays in RT-PCR returns.

**Figure 3:**
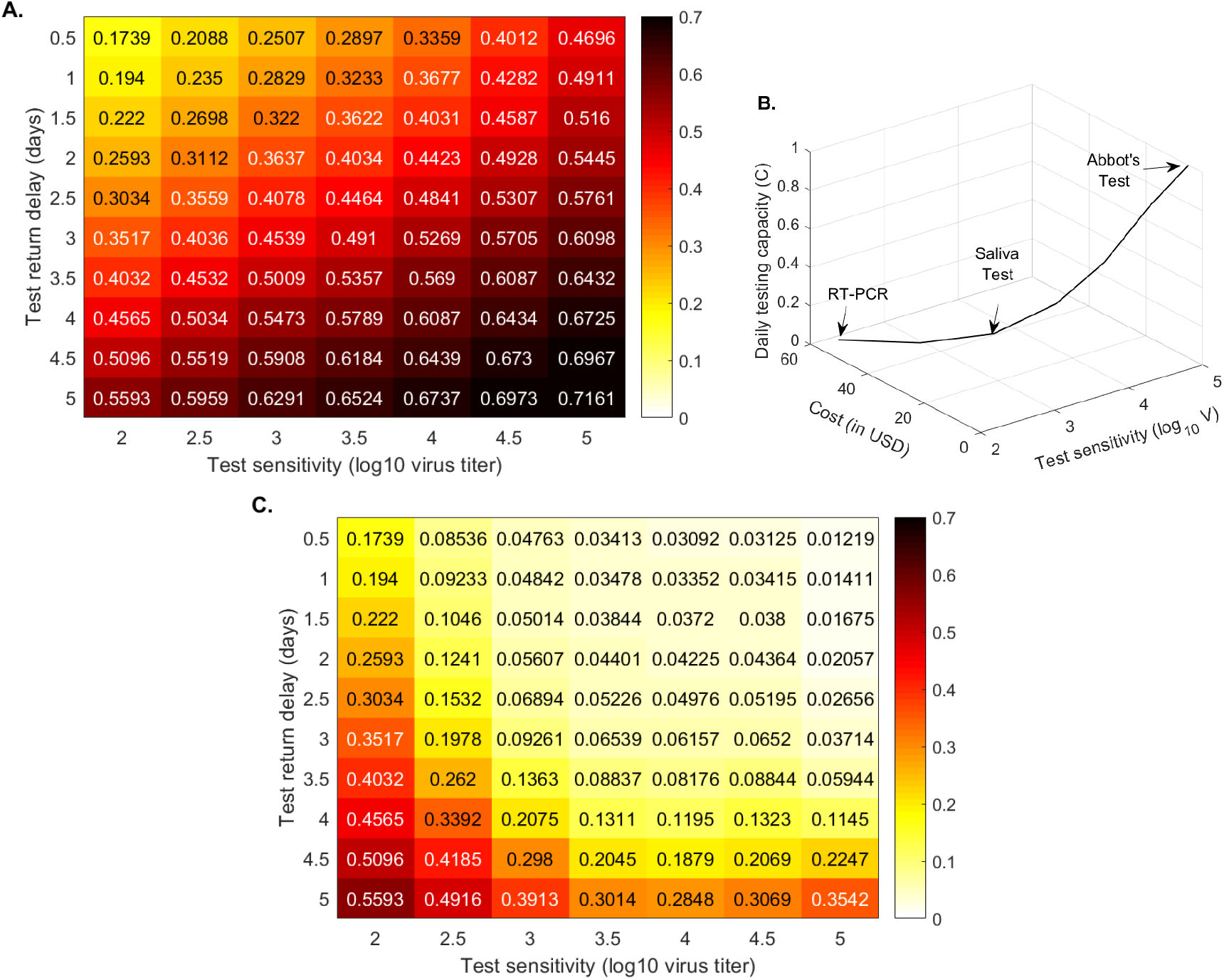
Cumulative cases at half a year. Heatmaps for the cumulative cases at half a year after the outbreak (% of the total population) as given by model eq. (5) versus test sensitivity and test return delay. Panel A: fixed testing capacity per day, *C* = 0.1. Panel B: relationship between capacity and cost. Panel C: fixed testing budget per day. Parameters and initial conditions are given in Tables 1 and 2.

### 3.4 Quantifying the tradeoff between test sensitivity and test frequency

We next investigate the effect that increased testing frequency has on the outcomes. While, under the Families First Coronavirus Response Act, testing in the United States is free of cost for an individual, the overall public health (or institutional) budget associated with test administration and processing may limit the overall number of tests available for administration each day. Conversely, reduction in test cost allows for increased testing capacity and frequency. We use model eq. (5) to quantify the overall infection, half a year after the start of the outbreak, when we provide as many tests as possible under a fixed daily budget.

Early and current studies show varied cost ranges for molecular and/or antigenic tests [26]. We assume the following costs for a single RT-PCR [26], Yale saliva [40], and Abott BinaxNow [1] tests: 50 USD, 10 USD and 5 USD, respectively. When we administer RT-PCR tests costing 50 USD, the daily testing capacity is equal to 10% of the population, *C* = 0.1, as in the previous sections. When we administer a saliva test costing 10 USD, the daily testing capacity is equal to 50% of the population, *C* = 0.5. Lastly, when we administer an Abott BinaxNow test costing 5 USD, the daily testing capacity is equal to 100% of the population, *C* = 1. Under these assumptions, the daily budget is the same regardless of the testing strategy. Moreover, we extrapolate these values to obtain intermediary cost functions (see Figure 3, panel B). We next derive a heatmap for the total cases, half a year after the start of the outbreak, for equal budget, varied testing sensitivities, and varied test return delays (see Figure 3, panel C).

We determine that tests of low-sensitivity (10^5^ virus load per swab detection and half a day return delay) that are administrated daily vastly outperform high-sensitivity tests. In particular, half a year after the start of the outbreak, the total number of cases is reduced from 19.4% for a RT-PCR that is returned in 24 hours (25.9% for a 48 hours return) to less than 1.2% when the low-sensitivity rapid test is given to everyone every day (see Figure 3, panel C). This is not a transient result, with overall infection reaching a maximum of 3.2% three years after the start of the outbreak. If the same low-sensitivity test is administered every other day, the overall infection is reduced to 3.8% half a year after the start of the outbreak; and if everyone is tested once every three days, the overall infection is reduced to 5.4% (not shown). If, however, everyone is tested with low-sensitivity tests once a week, than the overall infection is 30% half a year after the start of the outbreak (not shown), as high or higher than for the RT-PCR tests that are returned in less than 2.5 days. There is, therefore, a clear tradeoff between frequency, test sensitivity, and test return delays, which should be optimized to the needs of each community.

### 3.5 Transmission according to infection status

We investigate how testing regimes differentially affect the proportion of transmission associated with each disease status (symptomatic, presymptomatic and asymptomatic). We define presymptomatic, as infections that occur before day *τ*_*presym*_ = 4 days [18]. As seen in the previous sections, under fixed *C* = 0.1 daily testing, the peak daily incidence is reduced by 38.2% and 9.0%, respectively, when the antigen or paper-strip testing regimes replaced the standard RT-PCR tests. We further split the peak daily incidence into infections that occur due to symptomatic, presymptomatic and asymptomatic transmission (see Figure 4, orange vs red vs blue bars). Using the antigen test, peak daily incidence due to symptomatic transmission is reduced by 39.9% compared with RT-PCR, presymptomatic transmission is reduced by 33.2% and asymptomatic transmission is reduced by 38.2%. Using the paper-strip test, peak daily incidence due to symptomatic transmission is reduced by 8.9% compared with RT-PCR, presymptomatic transmission is reduced by 9.2% and asymptomatic transmission is reduced by 9.0% (see Figure 4, orange vs red vs blue bars).

**Figure 4:**
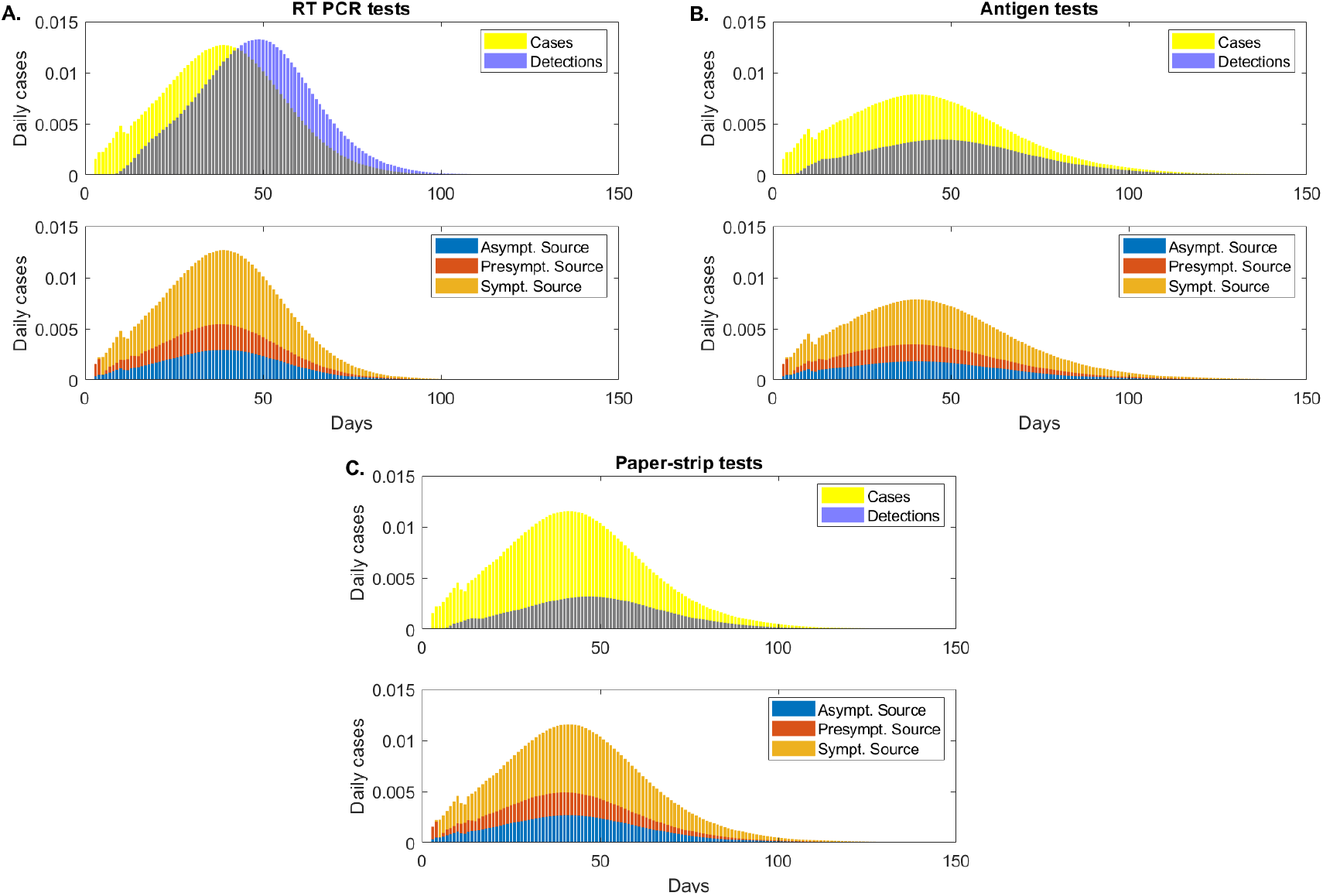
Asymptomatic, presymptomatic and symptomatic transmissions. (Upper figures): daily cases (yellow bars) and daily detections (blue bars); (Lower figures): daily cases due to asymptomatic transmission (blue bars), presymptomatic transmission (red bars) and symptomatic transmission (orange bars), as given by model eq. (5) for fixed testing capacity. Panel A: RT-PCR, detection threshold log_10_(*V*) = 2, test return delay 5 days; Panel B: antigen test, detection threshold log_10_(*V*) = 5, test return delay 0.5 days; Panel C: paper-strip test, detection threshold log_10_(*V*) = 6, test return delay 0.1 days.

This suggest that low-sensitivity tests are better than RT-PCR at reducing peak incidence in all types of transmissions, but the lower sensitivity paper-strip test shows limited improvement over the RT-PCR. Since the testing capacity is fixed, these results reflect the tradeoff between sensitivity and test return delays.

To account for the lower costs associated with antigen and paper-strip tests, we also calculate the reduction in peak daily incidence when these tests are administered at higher frequency than RT-PCR. As before, we assume that RT-PCR is administered at a fixed daily capacity of *C* = 0.1. When antigen tests are administered at daily capacity *C* = 0.3, *C* = 0.6 and *C* = 1 the total peak daily incidence is reduced by 70.4%, 76.2% and 79.0%, respectively. When paper-strip tests are administered at these capacities, the total peak daily incidence is reduced by 68.3%, 73.1% and 77.4%, respectively. We see limited variability in the reduction of infection due to symptomatic, presymptomatic and asymptomatic transmission (see Table 3).

**Table 3:** Percent reduction in daily incidence transmission for antigen and paper-strip tests at various daily testing capacities compared to daily incidence transmission for a RT-PCR test administered at *C* = 0.1 testing capacity per day.

| Test type | Infectious subgroup | $C = 0.1$ | $C = 0.3$ | $C = 0.6$ | $C = 1$ |
| --- | --- | --- | --- | --- | --- |
| Antigen | Symptomatic | 39.8% | 70.5% | 76.5% | 76.8% |
|  | Presymptomatic | 33.2% | 70.2% | 75.4% | 85.7% |
|  | Asymptomatic | 38.2% | 70.4% | 76.2% | 79.0% |
|  | Total | 38.2% | 70.4% | 76.2% | 79.1% |
| Paper-strip | Symptomatic | 8.9% | 68.4% | 74.0% | 76.8% |
|  | Presymptomatic | 9.2% | 68.2% | 70.5% | 79.3% |
|  | Asymptomatic | 9.0% | 68.3% | 73.1% | 77.4% |
|  | Total | 9.0% | 68.3% | 73.1% | 77.4% |

When antigen and paper-strip tests are administered with the same capacity as RT-PCR, *C* = 0.1, the antigen test significantly outperforms the paper-strip test in reducing peak daily incidence (38.2% vs. 9.0% reduction). However, as the capacity is increased, this difference in performance vanishes, and both tests approach a limiting peak incidence reduction of approximately 80% (see Table 3). This indicates that there is a critical capacity required to achieve significant incidence reductions with less sensitive tests.

## 4 Discussion

While RT-PCR is the gold-standard for diagnosis of SARS-CoV-2 cases, there are significant challenges in implementing effective epidemic surveillance and mitigation regimes on the basis of these tests, due to the need for specially trained lab personnel, limited lab capacity, high costs per test and delays in returning test results. Alternative tests such Abbott BinaxNOW^™^ can produce results rapidly, at lower cost and without the need for specialized lab personnel, but are less sensitive to lower virus concentrations. We investigated the tradeoffs between test sensitivity, return delay and test frequency using a deterministic mathematical model of virus transmission.

Our model shows that for fixed testing capacity, lower sensitivity tests with shorter return delays slightly flatten the daily incidence curve and delay the time to the peak daily incidence. The cumulative number of infections, however, shows a more complicated interaction between the loss of sensitivity and the benefits of faster test returns. We find that low-sensitivity tests with a return delay of one half day, such as antigen tests, reduce the cumulative case count at half a year into the outbreak. Despite the higher sensitivity of RT-PCR, in order to outperform the antigen test, its return delay would need to be reduced below 3 days. On the other hand, super-low-sensitivity tests with a return delay of 2 −3 hours, such as paper-strip tests, result in a cumulative case count slightly higher than RT-PCR.

The predicted mild improvement in cumulative case counts when low sensitivity tests replace to RT-PCR testing can be accentuated by increasing the total number of tests administered daily. Since antigen and other lower sensitivity tests are cheaper to produce and conduct, they can be delivered at higher frequency. We first varied testing capacity to account for the differing costs of tests, while keeping the testing budget fixed. This allows for daily testing of the entire population when antigen tests (which cost $5) are administered, but only 10% of the population when RT-PCR tests (which cost $50) are administrated. We found a large reduction in cumulative case counts, to as low as 1.5%, half a year into the outbreak and 3.2% three years into the outbreak. We next checked whether the magnitude of the improvement was preserved for lower daily antigen testing capacities. When antigen tests are administered to only 50% or 33% of the population daily, corresponding to testing the entire population every two or three days, respectively, we find that the decrease in the cumulative case counts persists. Thus, replacing RT-PCR testing with more frequent testing with less sensitive tests can lead to significantly improved outcomes, even if the testing budget is reduced by one half or one third.

We also studied whether there is a differential impact of alternative testing strategies on the proportion of viral transmission from sources at different stages of infection. When compared with RT-PCR, antigen and paper-strip tests reduce the number of new infections due to symptomatic, presymptomatic and asymptomatic sources by roughly equal amounts. This reduction is greater for increased testing frequency, however, the improvement is capped at approximately 80%.

Our modeling approach includes several simplifying assumptions, some of which can be relaxed to generalize our results in a variety of ways. First, we assume a well-mixed population and the model is therefore most suitable to a tightly interconnected community such as a college campus. Our findings support the conclusions of Paltiel, et al. [32], who found that frequent (every 2 days), low-sensitivity testing might be necessary in order to allow for college reopening. Moreover, several modeling studies have found that diagnostic testing of symptomatic patients alone is insufficient for outbreak control, and must be supplemented by randomized surveillance testing of the asymptomatic population [8, 11, 22, 32]. Indeed, under randomized and uniformly distributed surveillance testing of the entire non-isolated population, we find that frequent testing of the entire population can flatten the daily incidence curve and significantly decrease the cumulative size of the outbreak. Further work is needed to compare randomized testing to alternate strategies such as prioritizing the testing of high-risk or symptomatic individuals or preemptively quarantining those with symptoms and testing only asymptomatic individuals.

We have assumed a 100% detection rate when tests are administered to patients whose viral load is above the sensitivity threshold. As mentioned before, the BinaxNOW^™^ antigen rapid test has sensitivity levels of 85.7% for Ct< 25 (when the virus still infects), and 36.4% for Ct> 30 (when the virus may no longer be infectious) [22, 33]. We assume a step-function dependence of detection on viral load, with 0% detection below the threshold and 100% detection above. Moreover, we assume that all infected individuals have identical viral dynamics over the course of infection. Once available, more complete information about patient viral profiles and the dependence of test sensitivity on viral load can be incorporated to increase the accuracy of the model and to quantify the incidence of false negatives.

In summary, our study shows that surveillance testing that employs low-sensitivity tests at high frequency is an effective tool for epidemic control. Reduced cost per test is essential for the success of this approach, as it allows for the increased testing frequency, which overcomes sensitivity concerns. This more effective testing strategy would enhance the effectiveness of control measures that are testing-dependent, such as contact tracing, isolation and quarantining, further increasing our ability to overcome the COVID-19 epidemic.

## Data Availability

No original data is used in this modeling paper.

## Acknowledgments

SMC acknowledge funding from National Science Foundation grant No. 1813011.

## Author contributions

Conceptualization, J.E.F. and S.M.C.; programming, J.E.F.; writing, reviewing and editing, J.E.F. and S.M.C. All authors have read and agreed to the published version of the manuscript.

## Appendix

### Numerical scheme

The domain of model (5) is

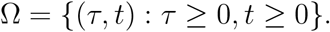

To numerically integrate system (5), we fix the ending time *T* and select a maximum age *G* greater than *T*. This allows us to find maximum values for 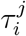, for *i*∈ *{*1,.., 4} and *j*∈ *{a, s}*, without losing any information. We construct a numerical scheme on the domain

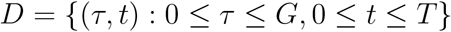

as follows. We discretize by taking equally spaced steps along the individual age of infection and the population time-since-outbreak, Δ*τ* = Δ*t*. Let *K* = ⌊*G/*Δ*τ* ⌋ and *Q* = ⌊*T/*Δ*t*⌋. Then, the age and time steps become *τ*_*k*_ = *k*Δ*t* and *t*_*q*_ = *q*Δ*t*, for 1 ≤ *k* ≤ *K* and 1 ≤ *q* ≤ *Q*. The delay *ℓ* will comprise *L* = ⌊*ℓ/*Δ*τ* ⌋ time steps.

### Initialization

We initialize the system with *S*^1^ = *S*(0) and

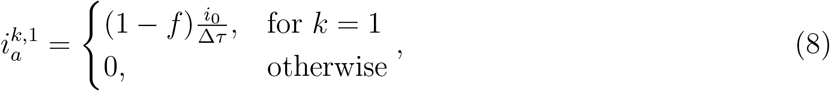

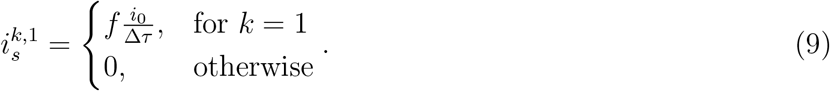

The initial infected population is assumed to have infection age *τ* = 0 at time *t* = 0, split between symptomatic and asymptomatic classes according to the ratio *f*. The total initial infected population is

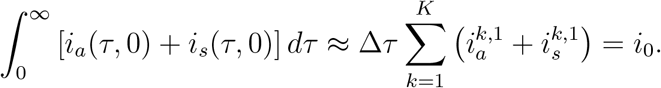

### Discretized Functions

Discretized versions of the functions *λ*_*a*_, *λ*_*s*_, *r*_*a*_ and *r*_*s*_ are needed. The force of infection terms *λ*_*a*_(*τ*) and *λ*_*s*_(*τ*) are independent of *t* and discretized versions are defined by

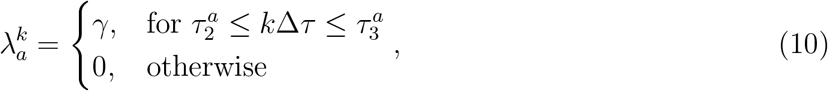

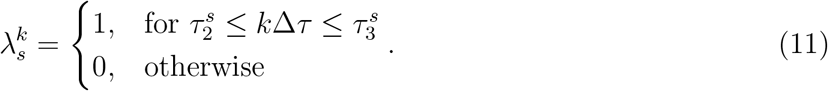

The testing rate *ρ*_rand_(*t*) depends on *t*, so at each time step *q* we calculate 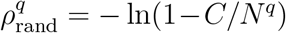, where *N*^*q*^ is the total current testable population

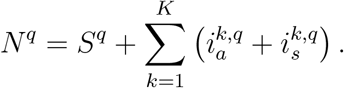

Let *j* ∈ {*a, s*} as appropriate. The discretized detection rates *r*_*j*_ are

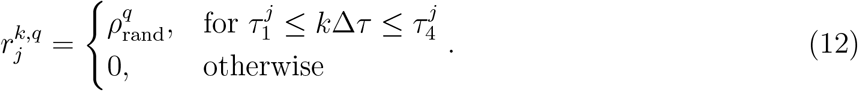

### Updating state variables

Given values for all state variables and all age classes at time step *q*, we update all state variables to time step *q* + 1. First, we calculate *i*_*a*_ and *i*_*s*_ at time step *q* + 1 for each age class except the first. For *k* ≤ *L* or *q* + 1 ≤ *L*, no positive test can have been returned, so *i*_*j*_ is governed by

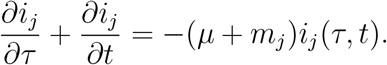

Using the method of characteristics, this equation can be solved precisely over the square [*k*Δ*t*, (*k* + 1)Δ*t*] × [*q*Δ*t*, (*q* + 1)Δ*t*] to give

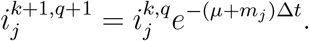

For *k* > *L* and *q* + 1 > *L*, testing and removal affects the dynamics of the infected classes, so *i*_*j*_are governed by

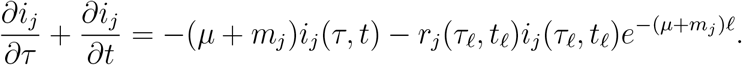

If we assume that the second term on the right hand side is a constant over the domain [*k*Δ*t*, (*k* + 1)Δ*t*] × [*q*Δ*t*, (*q* +1)Δ*t*], we can again use the method of characteristics to integrate over this square. This results in

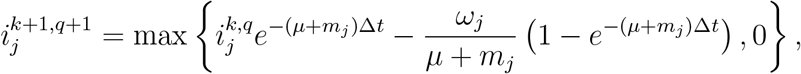

Where

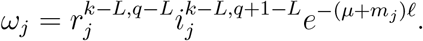

Next, we calculate the integral representing the force of infection.

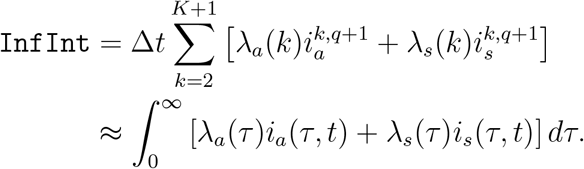

Third, we calculate the updated value of *S* using the standard implicit method

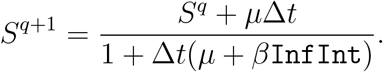

Finally, we fill in the age 0 infection level

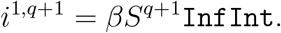

This completes the update of the scheme from time step *q* to time step *q* +1 for all state variables and all age classes.

